# Association of Intestinal Alkaline Phosphatase and vitamin D level with enteric pathogens in children: results from the MAL-ED study in Bangladesh

**DOI:** 10.1101/2025.10.01.25337051

**Authors:** Jafrin Ferdous, Md Amran Gazi, Md. Zahidul Islam, Rahvia Alam Sthity, Ishita Mostafa, Mustafa Mahfuz, Tahmeed Ahmed

## Abstract

**Objective:** Intestinal Alkaline Phosphatase (IAP), expressed by enterocytes, maintains gut homeostasis by promoting commensal microbiota growth and limiting inflammation. Vitamin D strengthens intestinal barrier, and its deficiency is linked to various diseases. This study explores the relationship among IAP, vitamin D, and enteric pathogens in children.

**Methods:** In the MAL-ED (Etiology, Risk Factors, and Interactions of Enteric Infections and Malnutrition and the Consequences for Child Health and Development) study, 265 newborns were enrolled between February 2010 to February 2012, from Dhaka, Bangladesh. 15-month time point data was collected and analyses, where intestinal pathogens were identified by TaqMan Array Card assay, stool IAP and serum vitamin D level were determined by colorimetric assay and chemiluminescence immunoassay respectively. Multivariable linear regression was performed to determine the association between IAP, vitamin D and gut pathogens. A p-value of <0.05 is considered as significant.

**Results:** Median (IQR) of IAP was 55.41 (19.60, 155.41) U/µL. Bivariate analysis showed significant negative association of IAP with *Campylobacter-pan Genome* (p=0.047), *Campylobacter jejuni* (p=0.020) and *Enterocytozoon bieneusi* (p=0.023). Multivariable linear regression analysis revealed a significant and inverse association with the *Adenovirus-pan genome* (β= -2.00; 95% CI= -3.94, -0.07) and *E. bieneusi* (β= -3.03; 95% CI= -5.94, -0.11).

**Conclusion:** IAP was inversely and significantly associated with *Adenovirus-pan genome* and *E. bieneusi,* a less-explored enteric pathogen, while its relationship with vitamin D was positive but not statistically significant. These findings suggest that IAP may play a protective role against specific enteric pathogens.

**What is already known on this topic:**

- Intestinal Alkaline Phosphatase (IAP) plays a crucial role in maintenance of gut homeostasis by promoting beneficial microbiota and reducing intestinal inflammation.
- Vitamin D strengthens the intestinal barrier, and deficiency has been linked to increased risk of infections and various chronic diseases.
- While both IAP and vitamin D are important in gut health, evidence linking them with specific enteric pathogens in human, especially in children remains limited.

**What this study adds:**

- This study demonstrates that IAP is inversely and significantly associated with *Adenovirus-pan genome* and *Enterocytozoon bieneusi*, a less-explored pathogen in children.
- It shows that although not statistically significant, IAP levels were positively related to vitamin D
- These findings provide novel evidence that IAP may play a protective role against specific viral and protozoan enteric infections.

**How this study might affect research, practice or policy:**

- The results highlight IAP as a potential biomarker of intestinal defense and a candidate for future therapeutic or preventive interventions targeting childhood enteric infections.
- Insights into IAP–pathogen relationships could guide in the development of microbiome-based or nutritional strategies to reduce diarrheal disease burden, and support the inclusion of IAP as potential intervention in child health and nutrition research in low-resource settings.

## Introduction

Alkaline phosphatases (AP) are enzymes that hydrolyze monophosphate esters by removing their phosphate components. The two primary categories of AP are tissue-specific alkaline phosphatases, which include germ cell alkaline phosphatase, placental alkaline phosphatase, and intestinal alkaline phosphatase (IAP); and tissue-non-specific AP that are present in bone, liver, and kidney ^1^. The luminal vesicles in the brush border membrane of intestinal epithelial cells are the site of elevated expression of IAP at a favourable pH of around 9.7. The main site of IAP secretion is the duodenum, with a lesser amount in the jejunum, ileum, and colon; the stomach is devoid of any IAP ^2–4^. Major elements of Gram-negative bacterial cell walls are lipopolysaccharides (LPS), the atypical presence of which in blood causes strong inflammatory responses in cases like sepsis and also chronic inflammatory responses in obesity, heart disease, and diabetes mellitus. IAP plays a vital role in preventing bacterial endotoxin-induced inflammation or sepsis by dephosphorylating the endotoxin-containing LPS, thus acting as a gut mucosal defence factor ^1^ ^4^. IAP deficiency has been linked to plethora of conditions like many gastrointestinal disorders-Inflammatory Bowel Disease (IBD), necrotizing enterocolitis; type 2 diabetes, ischemic heart disease, aging, and metabolic syndrome ^5^ ^6^ and exogenous IAP supplementation improves the outcomes linked to these disorders ^7^. The extensive functions of IAP encompass: intestinal epithelial barrier protection by modulating the levels of tight junction proteins ^8^, the regulation of gut microbial communities via antimicrobial proteins like lysozymes that control bacterial populations in the intestine ^9^. Thus, IAP plays a crucial role in promoting mucosal immunity, providing protection against enteric pathogens, maintaining the harmony of the gut tract by regulating intestinal epithelial cell turnover ^10^.

The human body is home to approximately 10-100 trillion microbial cells maintaining a symbiotic relationship, the gut being the main site for them due to its warm, stable, and eutrophic environment. These microbial cells are called the gut microbiome ^11^, the dysbiosis of which causes significant gastrointestinal symptoms such as mild to severe gastroenteritis, chronic diarrhea, malabsorption, and wasting. *Enterocytozoon bieneusi* is a not-so-common human pathogen, the epidemiology of which is still not very clear despite scientific advances. Since its first identification as an opportunistic pathogen related to human acquired immunodeficiency syndrome (AIDS), numerous cases of chronic diarrhea and intestinal microsporidiosis caused by this microorganism have been documented in those with compromised immune systems (such as recipients of organ transplant), as well as in healthy travelers, children, and in elderly. Immunocompromised individuals have also been observed to have hepatobiliary and pulmonary involvement ^12^.

Conversely, non-enveloped, icosahedral, double-stranded DNA viruses known as *Adenoviruses* (AdVs) are frequently found in vertebrate host species which can cause mild to severe illnesses in humans and other primates, including gastroenteritis, epidemic conjunctivitis, and respiratory infections ^13^. As seen in viral gastroenteritis, the disruption of epithelial cells caused by *Human Adenoviruses* (HAdV) may indirectly upset the equilibrium of the gut microbiota, causing or exacerbating diarrhea ^14^. HAdV cause severe diarrhea and are linked to high morbidities and mortality rates in both adult and children, the latter having a higher prevalence of HAdV infection due to their underdeveloped immune systems ^15^ ^16^. HAdVs pan-genome refers to the aggregate of all genomes in a clade and is divided into seven species (A–G), which currently encompasses 70 types. *Adenovirus F*, which represents *Adenovirus types 40* and *41* (AdV-40, AdV-41), has been identified as a subgenus that is associated with acute gastroenteritis, are prevalent year-round and primarily affect minors under the age of 2 ^17^ .

Vitamin D signaling has been demonstrated to improve gut barrier function by upregulating the expression of tight junction proteins (TJP) such as Zonula Occludens-1 and Occludin in the gut, as well as influencing the gut microbiota ^18^. Vitamin D is acquired from production by skin exposed to sunlight and also from dietary sources and it’s deficiency has been linked to conditions such as Inflammatory Bowel Disease (IBD), diabetes, cancer, and autoimmune diseases etc. and is common in children ^19^. Vitamin D, being a common inducer, can upregulate IAP activity, which is crucial for the maintenance of intestinal homeostasis ^7^. Vitamin D was found to enhance the transcriptional expression of IAP and it’s activity in Caco-2 cells in colorectal adenocarcinoma patient ^20^. Rats on a high-fat diet with vitamin D restriction also showed decreased IAP activity in their duodenum ^7^. These results lend credence to the hypothesis that IAP expressions in gastrointestinal homeostasis may be significantly influenced by an adequate vitamin D intake ^21^.

A critical context where these interplays become especially relevant is Environmental Enteric Dysfunction (EED), a common subclinical condition among children in low- and middle-income countries. EED, a widespread disruption of small intestinal structure and function ^22^, has been closely linked to growth and developmental delays, poor oral vaccine efficacy, and growth failure ^23^ ^24^. The aetiology is multifactorial, driven by recurrent enteric infections, poor sanitation, and gut-immune dysregulation precisely where IAP and vitamin D signalling play key roles. IAP’s anti-inflammatory and barrier-protective effects suggest its deficiency may contribute to EED pathogenesis ^25^, while its restoration presents a potential therapeutic strategy ^26^. We aim to evaluate the potential correlation between enteric pathogens, vitamin D, and IAP in our analysis of MAL-ED participants ^27^ ^28^.

## Methods

### Study Population, Timeline, Settings and Sample Size

The MAL-ED study was conducted in 8 countries in Asia, Africa and South America; namely: Bangladesh, India, Pakistan, Nepal, Brazil, Peru, South Africa, and Tanzania. Here, we’ve analysed data from only Bangladesh. A census was conducted in Bauniabadh slum area of Mirpur, Dhaka, Bangladesh to assess the under-5 children number and the reproductive age group women number. This data helped the site to determine the catchment area where the estimated number of infants would be born within the 2 years enrolment period. The Bauniabadh area in Mirpur, Bangladesh is a crucial location for contributing to the study because of the high prevalence of gastrointestinal infectious morbidity, malnutrition, inadequate sanitation, and high population density ^28^. The MAL-ED study had 3 components, the birth cohort component being one of them. In this component, 265 healthy newborn children were enrolled within their 1^st^ week of birth, between February 2010 to February 2012, from an urban slum of Bauniabadh, Mirpur. 80% of the total recruitment, that is 211 of the children, were followed up for 24 months ^29^. The enrolled infants were </= 17 days of age with a birth weight of >1500g ^30^. Children with congenital defects or any severe disease (i.e., Cleft lip or palate, Cerebral palsy, Chromosomal disorders, etc.) had been excluded. Details methodology of the study have been published previously ^28^.

The participants’ levels of intestinal pathogens, serum vitamin D, and IAP have been considered in this analysis. Cross-sectional data of a 15-month time point from the participants were collected and analysed ^31^. Intestinal pathogens were identified by TaqMan array card assay and the level of IAP in stool was determined by colorimetric assay technique. To detect the presence of 29 pathogens from each of the samples, TaqMan array cards, customized multiplex quantitative polymerase chain reaction involving compartmentalized primer-probe assays were used, the details are described elsewhere ^32^ ^33^. Stool samples were weighted and ddH_2_O (double-distilled water) were added at a defined ratio. Generally, 1 mg of stool was mixed with 50 μl of stool dilution. In order to prepare a homogenized stool mixture, samples were vigorously vortexed, centrifuged at 10,000 ×g for 20 min, and the supernatant comprising IAP was separated. The activity of IAP was determined by Alkaline Phosphatase Di-ethanolamine Activity Kit following the manuals provided with the kit (Sigma-Aldrich, USA). The stool IAP values are shown as units of IAP/g stool. Scheduled blood samples were collected at the 15-month time point, and serum vitamin D was measured by chemiluminescence immunoassay. Serum vitamin D concentration <50 ng/mL was defined as low serum vitamin D ^34–36^.

### Statistical Analyses

Bivariate analysis of the data revealed few significant associations between IAP and some notorious pathogens. Multivariate linear regression was performed further to determine the actual association between IAP, vitamin D and potential gut pathogens.

For categorical data, the baseline characteristics were represented by frequency and percentage; for asymmetric continuous data, by median with inter-quartile ranges (IQR); whereas for symmetric continuous variables, by mean with standard deviation. The t-test or Mann-Whitney U test, if appropriate, was used to assess the differences in IAP levels of the E. coli pathotype positive and negative groups. The IAP levels were square root transformed and made symmetric before being included in the multivariate model. Multivariable linear regression was applied to analyse the association between E. coli pathotypes, vitamin D, and IAP concentrations. All the models were adjusted by sex, total energy, myeloperoxidase, alpha-1 antitrypsin and minimum meal frequency (MMF-the proportion of breastfed children aged 9-23 month who received complementary foods three times, with at least one feed comprising solid, semi-solid, or soft food), which were selected on the basis of existing literature also those are biologically as well as clinically important. Separate models have been run for each of the pathogens. A multivariable linear regression was performed to determine this association, in which the IAP level was the outcome variable and the pathogens were the exposure variables. A probability of <0.05 was considered as statistically significant. Infant and young child feeding practices were assessed according to WHO/ UNICEF indicators. All analyses were performed in the statistical software STATA version 15.0 (StataCorp, College Station, Texas, USA).

## Results

211 of our enrolled participants have completed follow up and IAP data was available for 188 of them. For this analysis, 188 available samples from MAL-ED study were considered where 52% were female and 48% were male, average age of participants were 15 months. 24% participants had 1 episode and 3% had 2 episodes of diarrhoea, 6% had acute lower respiratory tract infections. The median IAP level was 55.4 with IQR (19.6, 155.4), exclusive breast-feeding duration was 105 days with IQR (57, 154), minimum dietary diversity (MDD-the proportion of children aged 6–23 months who consumed foods from at least five of eight recommended groups in the preceding 24 hours: breastmilk; grains, roots, tubers and plantains; legumes, nuts and seeds; dairy products; flesh foods; eggs; vitamin A-rich fruits and vegetables; and other fruits and vegetables) was present in 73% of the participants (table 1).

**Table 1:** Baseline characteristics of the participants in MAL-ED birth cohort.

| Characteristics |  | Total (188) |
| --- | --- | --- |
| Participant's sex, n (%) | Male | 90 (48) |
|  | Female | 98 (52) |
| Age in months, Mean (SD) |  | 15.0 (0.07) |
| Number of episodes of diarrhea n (%) | No episodes | 138 (73) |
|  | 1 Episodes | 45 (24) |
|  | 2 Episodes | 5 (3) |
| Acute Lower Respiratory Tract Infection n (%) | Yes | 11 (6) |
|  | No | 177 (94) |
| Number of episodes of cough n (%) | No episodes | 64 (34) |
|  | 1 Episodes | 94 (50) |
|  | 2 Episodes | 28 (15) |
|  | 3 Episodes | 1 (0.5) |
|  | 4 Episodes | 1 (0.5) |
| Number of episodes of fever n (%) | No episodes | 86 (46) |
|  | 1 Episodes | 78 (41) |
|  | 2 Episodes | 21 (11) |
|  | 3 Episodes | 2 (1) |
|  | 4 Episodes | 1 (0.5) |
| Maternal height in cm, Mean (SD) |  | 149.08 (4.98) |
| Participant's birth weight in Kg, Mean (SD) |  | 2.80 (0.42) |
| Duration of exclusive breast feeding in days, Median |  | 105 (57, 154) |
| Total Energy in Kcal, Median (IQR) |  | 272.99 (183.76, |
| Minimum Dietary Diversity n (%) | Present | 136 (73) |
|  | Absent | 51 (27) |
| Participant's birth length in cm, Mean (SD) |  | 48.20 (1.98) |
| Concentration of Myeloperoxidase, Median (IQR) |  | 3012 (1193, 5718) |
| Concentration of Neopterin, Median (IQR) |  | 1311 (454, 2750) |
| Concentration of Alpha-1 Antitrypsin, Median (IQR) |  | 0.31 (0.17, 0.58) |
| Intestinal Alkaline Phosphatase (IAP) level, Median |  | 55.4 (19.6, 155.4) |
| Stunting, n (%) | Present | 79 (42) |
|  | Absent | 109 (58) |

In this study, we have observed the prevalence of multiple pathogens in the gut of 15-month-old participants, where Enteroaggregative *Escherichia coli* had the highest prevalence (52%), followed by Enterotoxigenic *Escherichia coli* and *Adenovirus-pan genome* (46%) and *Campylobacter-pan genome* (44%). All other species, *Campylobacter jejuni* (37%), Atypical Enteropathogenic *E. coli* (aEPEC 33%), Heat Stable Enterotoxigenic *E. coli*, Typical Enteropathogenic *E. coli* (tEPEC 21%), Heat Labile Enterotoxigenic *E. coli*, *Bacteroides fragilis*, Fungus-*Enterocytozoon bieneusi* (13%) etc. had a prevalence at or below 40%. The details of prevalence are listed in the **figure 1**.

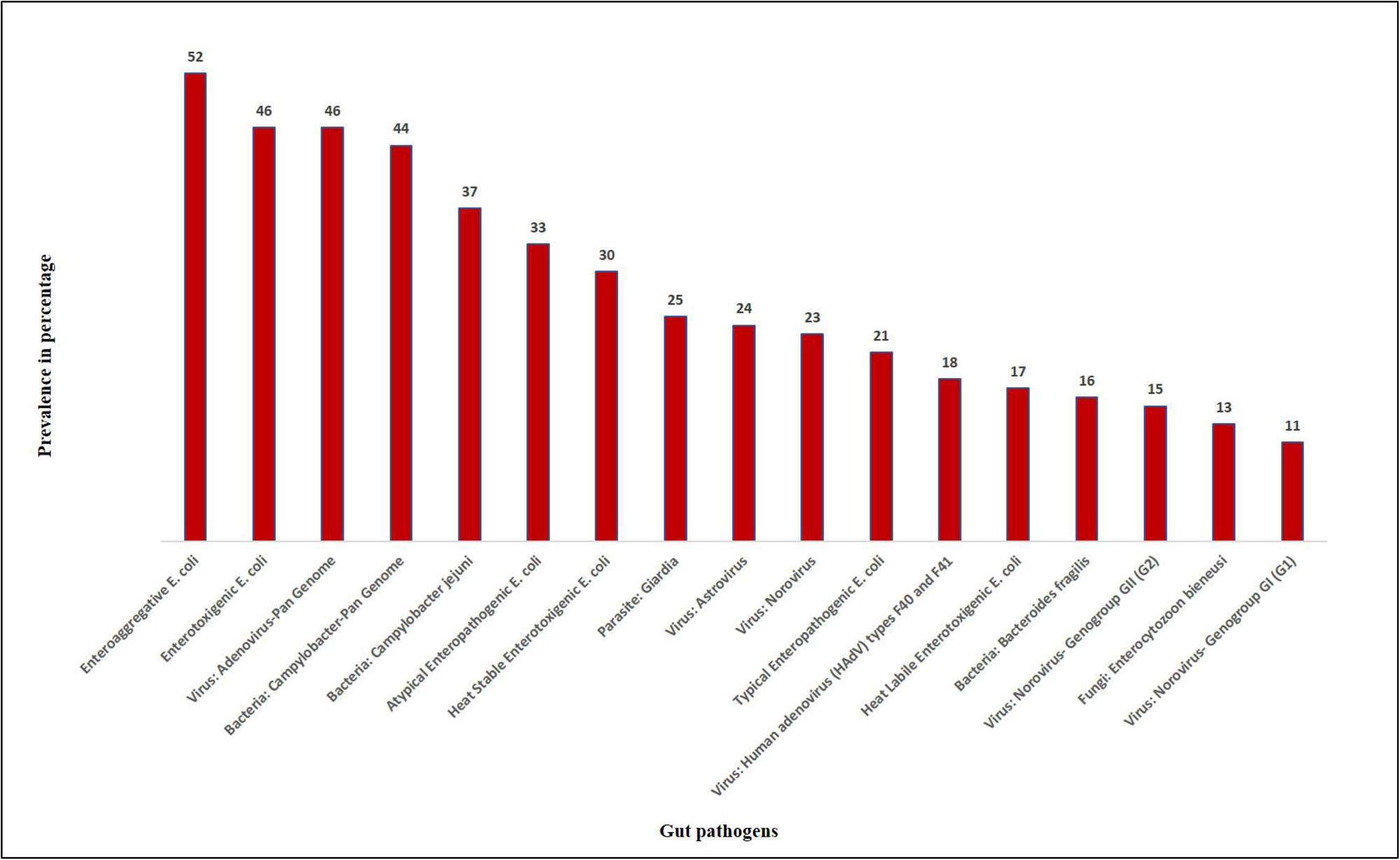

In bi-variate analysis, a significant association of IAP with *Campylobacter-pan genome* (p=0.047), *Campylobacter jejuni* (p=0.020), and *Enterocytozoon bieneusi* (p=0.023) was observed (table 2).

**Table 2:** Summary of IAP level of MAL-ED birth cohort by pathogens.

| Pathogens |  | IAP |  |
| --- | --- | --- | --- |
|  |  | Median (IQR) | p-value |
| Enteroaggregative *Escherichia coli* (EAEC) | Present | 50.7 (16.9, 153.4) | 0.82 |
|  | Absent | 52.7 (21.6, 152.7) |  |
| Enteropathogenic *Escherichia coli* (aEPEC) | Present | 50.7 (19.6, 189.2) | 0.72 |
|  | Absent | 52.7 (18.9, 129.7) |  |
| *Adenovirus-pan* genome | Present | 41.9 (18.9, 120.3) | 0.16 |
|  | Absent | 70.3 (21.6, 195.3) |  |
| ***Campylobacter-pan* genome** | **Present** | **35.1 (12.2, 127.0)** | **0.04** |
|  | **Absent** | **66.2 (22.9, 175.7)** |  |
| ***Campylobacter jejuni*** | **Present** | **31.8 (12.2, 127.0)** | **0.02** |
|  | **Absent** | **74.3 (22.9, 156.8)** |  |
| Enterotoxigenic *Escherichia coli* (ETEC) | Present | 42.6 (22.9, 129.7) | 0.66 |
|  | Absent | 52.7 (17.6, 154.1) |  |
| Heat Stable Enterotoxigenic *Escherichia coli* (ST_ETEC) | Present | 35.8 (14.9, 129.7) | 0.31 |
|  | Absent | 58.1 (18.9, 154.1) |  |
| *Giardia* | Present | 32.4 (14.9, 152.7) | 0.28 |
|  | Absent | 55.4 (22.9, 156.8) |  |
| *Astrovirus* | Present | 52.7 (21.6, 168.9) | 0.67 |
|  | Absent | 52.7 (18.9, 131.1) |  |
| *Norovirus* | Present | 41.9 (18.9, 129.7) | 0.83 |
|  | Absent | 52.7 (18.9, 154.1) |  |
| Typical Enteropathogenic *Escherichia coli* (tEPEC) | Present | 37.8 (18.9, 127.0) | 0.45 |
|  | Absent | 60.1 (18.9, 159.5) |  |
| *Human adenovirus* (HAdV) types *F40* and *F41* | Present | 51.4 (14.9, 110.8) | 0.34 |
|  | Absent | 50.7 (20.3, 158.1) |  |
| Heat Labile Enterotoxigenic *Escherichia coli* (LT_ETEC) | Present | 50.7 (22.9, 123.6) | 0.69 |
|  | Absent | 52.7 (17.6, 154.1) |  |
| *Bacteroides fragilis* | Present | 55.4 (22.3, 126.4) | 0.93 |
|  | Absent | 50.7 (18.9, 154.1) |  |
| *Norovirus- Genogroup GII* (*G2*) | Present | 33.8 (13.5, 125.7) | 0.57 |
|  | Absent | 52.7 (19.6, 156.8) |  |
| ***Enterocytozoon bieneusi*** | **Present** | **22.9 (9.5, 83.8)** | **0.02** |
|  | **Absent** | **58.1 (21.6, 160.8)** |  |
| *Norovirus- Genogroup GI* (*G1*) | Present | 58.1 (22.9, 156.8) | 0.78 |
|  | Absent | 52.7 (17.6, 152.7) |  |

**Figure 2** illustrates the correlation between intestinal pathogens and IAP.

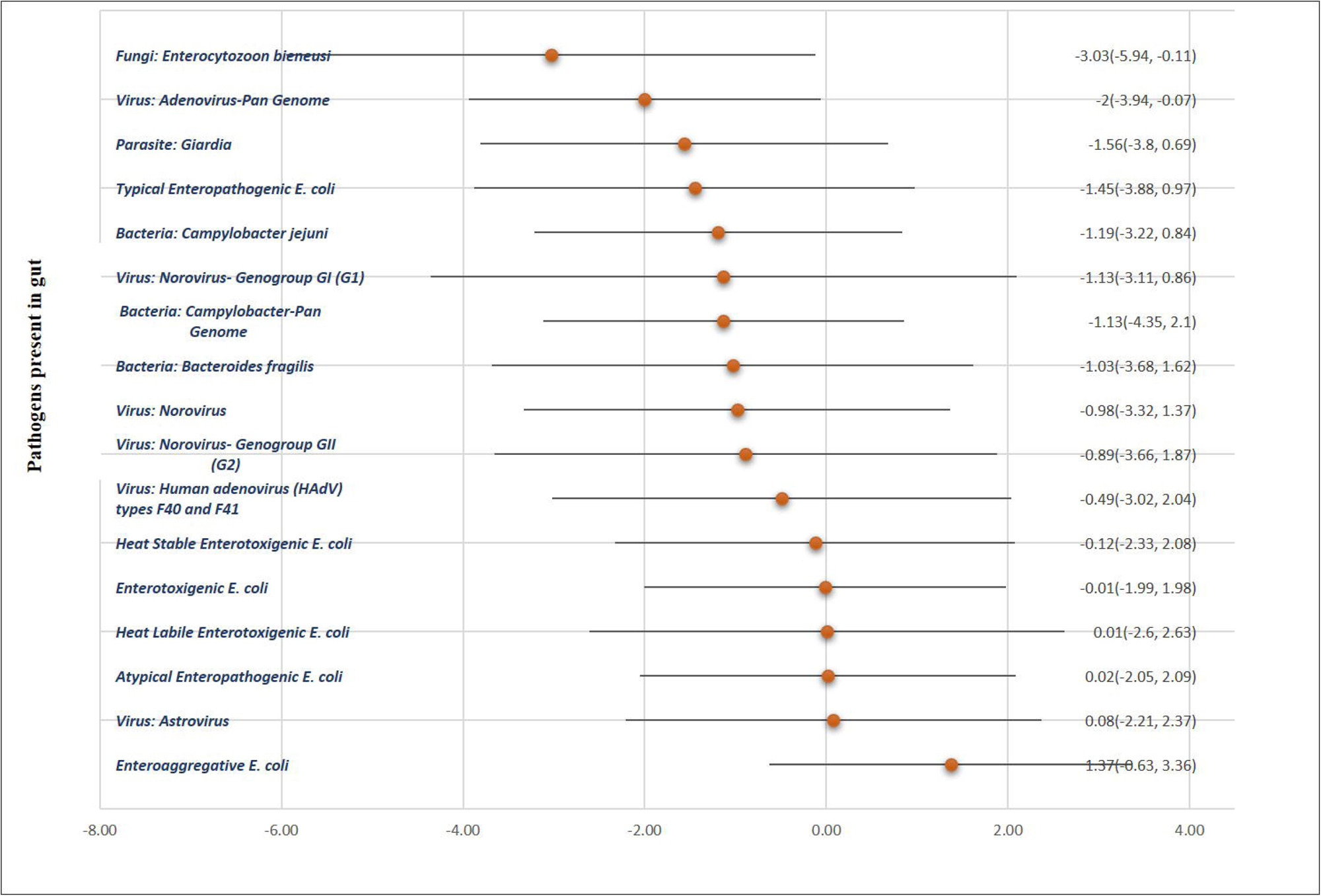

The summary of IAP and vitamin D by pathogens is shown in the tables-2, 3 and 4. The tables show that Bacteria: *Campylobacter-Pan Genome*, Bacteria: *Campylobacter jejuni* and Fungi: *Enterocytozoon bieneusi* were significantly associated with IAP, where in presence of those pathogens, the level of IAP was lower. In the multivariable linear regression analysis (table 4), the main exposure variable, IAP level, was positively associated with vitamin D concentration (β = 0.33, 95% CI: -0.16 to 0.81); however, this association was not statistically significant (p= 0.182). Although some variables such as EAEC positivity and *Giardia* presence showed positive coefficients, and others like female gender and minimum meal frequency showed negative associations, all confidence intervals included zero and p-values exceeded 0.05. These findings suggest that, within this model, neither IAP level nor the adjusted covariates were significantly associated with vitamin D concentration.

**Table 3:** Summary of serum vitamin D level of MAL-ED birth cohort by pathogens.

| Pathogens |  | Vitamin D |  |
| --- | --- | --- | --- |
|  |  | Median (IQR) | p-value |
| Enteroaggregative *Escherichia coli* (EAEC) | Present | 52.7 (43.1, 62.3) | 0.16 |
|  | Absent | 49.7 (39.3, 57.0) |  |
| Enteropathogenic *Escherichia coli* (aEPEC) | Present | 54.1 (43.0, 62.3) | 0.32 |
|  | Absent | 49.4 (40.5, 60.7) |  |
| *Adenovirus-pan genome* | Present | 53.0 (43.9, 62.3) | 0.38 |
|  | Absent | 50.6 (40.4, 59.9) |  |
| *Campylobacter-pan genome* | Present | 52.3 (43.6, 59.6) | 0.61 |
|  | Absent | 50.3 (39.9, 61.5) |  |
| *Campylobacter jejuni* | Present | 52.3 (43.0, 59.6) | 0.79 |
|  | Absent | 50.3 (41.2, 61.5) |  |
| Enterotoxigenic *Escherichia coli* (ETEC) | Present | 50.3 (42.4, 62.3) | 0.91 |
|  | Absent | 51.4 (41.4, 61.5) |  |
| Heat Stable Enterotoxigenic *Escherichia coli* (ST_ETEC) | Present | 48.2 (39.3, 62.3) | 0.79 |
|  | Absent | 51.4 (41.6, 60.7) |  |
| *Giardia* | Present | 49.2 (39.8, 58.0) | 0.55 |
|  | Absent | 52.5 (42.4, 61.5) |  |
| *Astrovirus* | Present | 48.2 (42.4, 54.0) | 0.10 |
|  | Absent | 52.7 (41.4, 63.9) |  |
| *Norovirus* | Present | 50.9 (43.1, 58.6) | 0.91 |
|  | Absent | 51.4 (41.2, 62.8) |  |
| Typical Enteropathogenic *Escherichia coli* (tEPEC) | Present | 49.5 (37.3, 60.3) | 0.53 |
|  | Absent | 51.7 (41.7, 61.5) |  |
| *Human adenovirus* (HAdV) types F40 and F41 | Present | 46.3 (42.1, 57.9) | 0.32 |
|  | Absent | 52.3 (41.6, 61.6) |  |
| Heat Labile Enterotoxigenic *Escherichia coli* (LT_ETEC) | Present | 51.2 (41.7, 56.8) | 0.92 |
|  | Absent | 51.2 (41.5, 61.1) |  |
| *Bacteroides fragilis* | Present | 54.0 (43.0, 57.0) | 0.98 |
|  | Absent | 50.6 (41.6, 62.3) |  |
| *Norovirus- Genogroup GII (G2)* | Present | 52.8 (43.6, 58.9) | 0.87 |
|  | Absent | 50.4 (41.3, 61.6) |  |
| *Enterocytozoon bieneusi* | Present | 56.2 (43.1, 69.6) | 0.21 |
|  | Absent | 49.9 (41.4, 58.9) |  |
| *Norovirus- Genogroup GI (G1)* | Present | 53.0 (45.5, 59.9) | 0.55 |
|  | Absent | 50.3 (41.4, 61.5) |  |

**Table 4:**
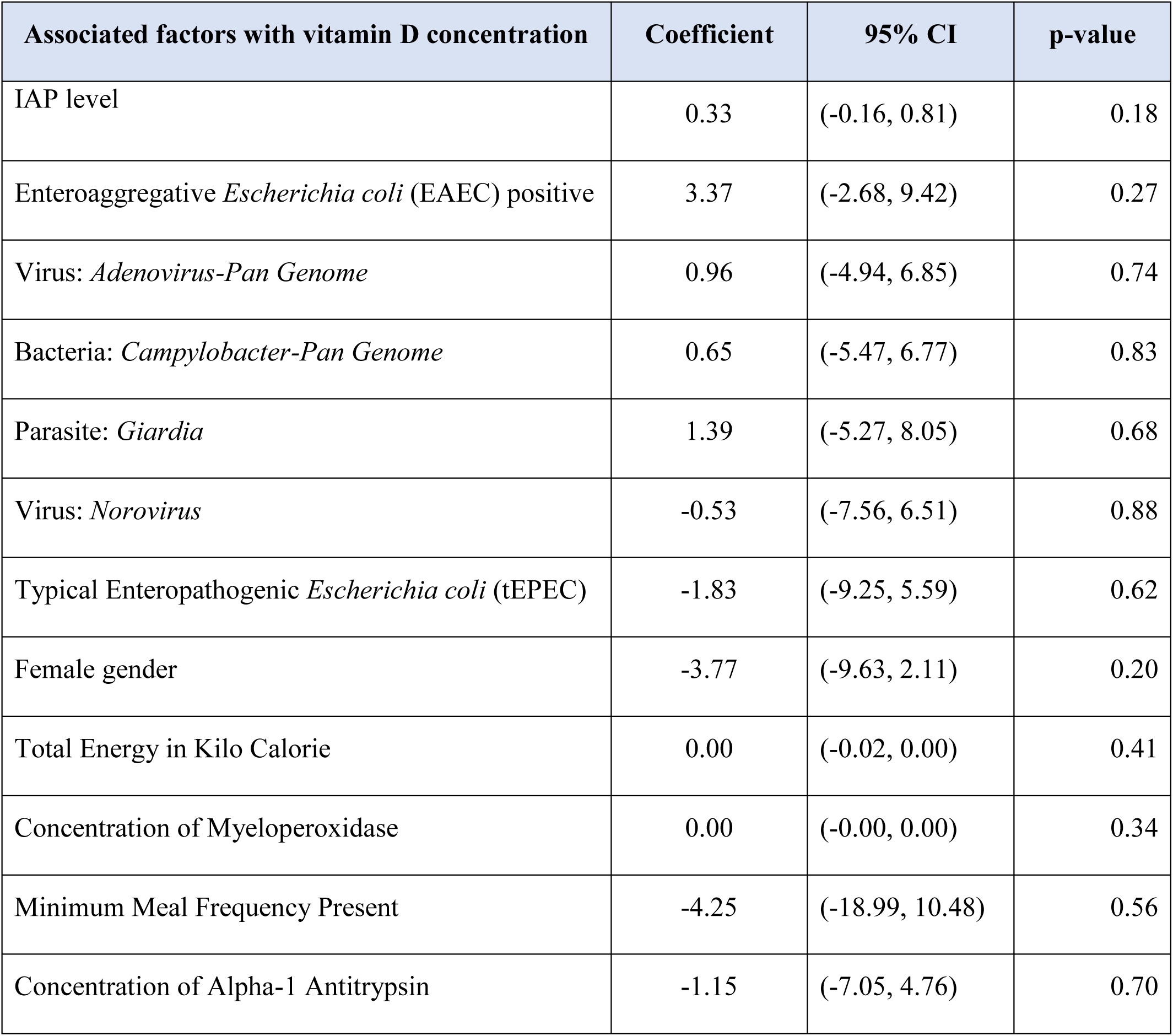
Associated factors of serum vitamin D level in 15-month old MAL-ED birth cohort.

Despite the fact that a significant association of IAP was observed with *Campylobacter-pan genome*, *Campylobacter jejuni*, and *Enterocytozoon bieneusi* in bivariate analysis, additional analysis revealed new findings. Multivariable linear regression analysis, however, revealed no significant association between IAP and *Campylobacter-pan genome* or *Campylobacter jejuni*, but showed a significant and inverse association with the *Adenovirus-pan genome* (β= -2.00; 95% CI= -3.94, -0.07; p= 0.043) as well as the *Enterocytozoon bieneusi* (β= -3.03; 95% CI= -5.94, -0.11; p= 0.042) (table 5).

**Table 5:**
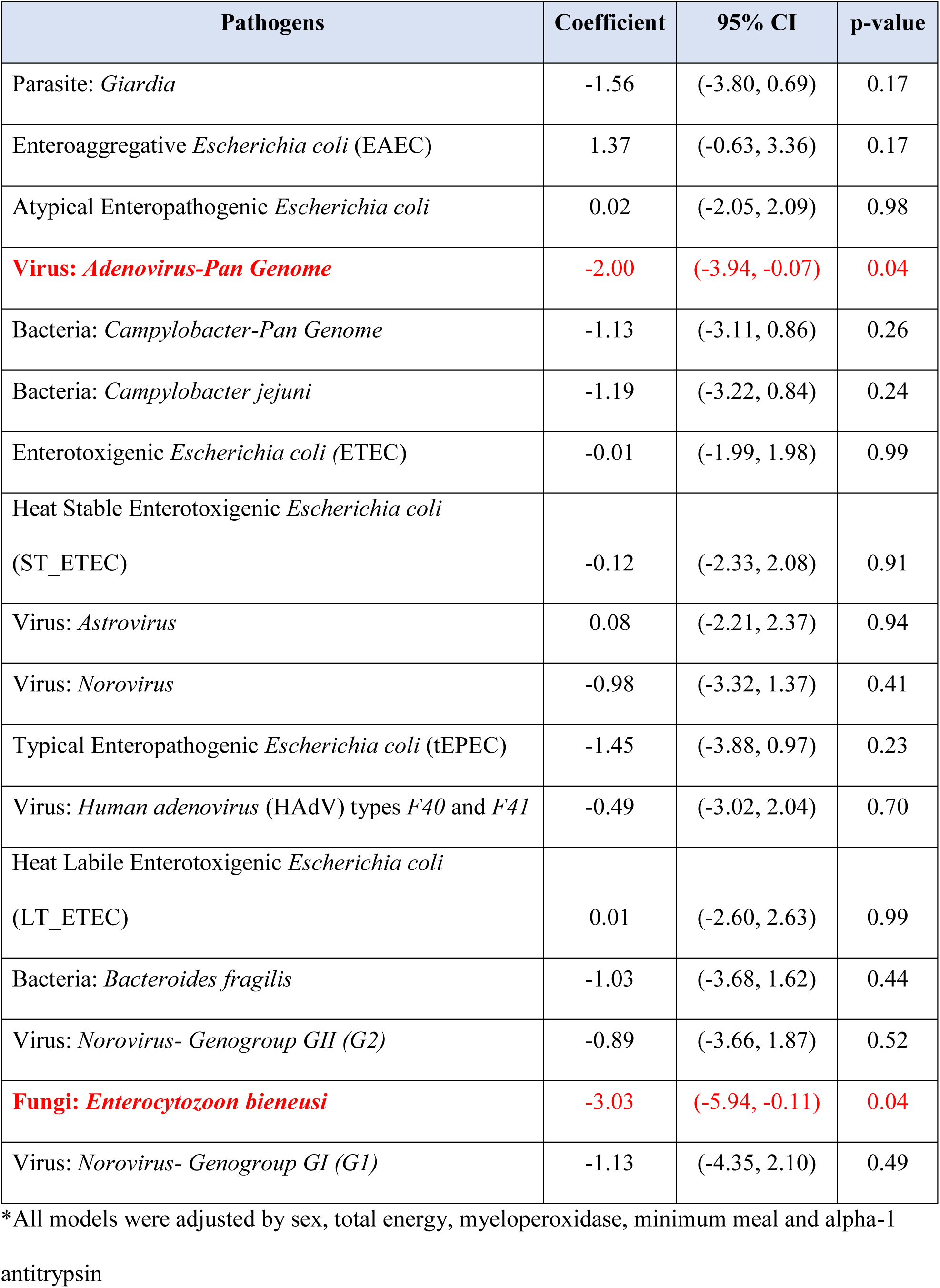
Results from association of IAP level with gut pathogens in MAL-ED birth cohort.

## Discussion

This study showed a novel perspective into the interplay between intestinal alkaline phosphatase (IAP), vitamin D levels, and numerous enteric pathogens, particularly *Enterocytozoon bieneusi* and *Adenovirus*. Our results point to a potentially significant axis of host defence control in the gut by indicating that lower IAP expression is linked to an increased incidence of enteric infections.

Studies showed that, IAP plays a crucial role in maintaining intestinal homeostasis by dephosphorylating bacterial products like Lipopolysaccharide (LPS) and ATP, thereby reducing inflammation and promoting a balanced gut microbiota and also its ability to support tight junction integrity and reduce bacterial translocation ^25^. In another study of vitamin D treatment in the chick intestinal mucosa, vitamin D-deficient chicks’ Intestinal alkaline phosphatase increases 2-3-fold after oral administration of 500 I.U. of vitamin D (cholecalciferol). Maximum IAP activity occurs 60 h following vitamin delivery. After oral vitamin D administration, calcium transport also increased in vitro across intact segments of ileal tissue, suggesting that IAP plays a role in vitamin D-mediated calcium transport. This study showed that vitamin D increases IAP activity and IAP in turn plays a pivotal role in vitamin D mediated calcium transport ^37^. Another study demonstrated that individuals with low serum vitamin D levels exhibited significantly reduced IAP activity, supporting the idea that vitamin D regulates IAP expression which suggests that vitamin D supplementation could indirectly boost IAP activity, enhancing mucosal immunity by improving both enzymatic and epithelial defence mechanisms ^38^.

*E. bieneusi* is an obligate intracellular pathogen affecting the intestinal epithelium, mostly in immunocompromised individuals as well as in healthy individuals. Our findings show an inverse association between IAP levels and *E. bieneusi* infection. Previous genomic studies have shown that *E. bieneusi* possesses a minimal yet highly efficient genome, with conserved virulence factors capable of modulating host immune responses ^39^. This supports the notion that *E. bieneusi* may actively or passively suppress IAP to establish infection. Vitamin D deficiency exacerbates this scenario more by weakening innate immunity and antimicrobial responses, which may explain why patients with both low vitamin D and reduced IAP had higher pathogen burdens ^40^.

Enteric *Adenoviruses* (especially species F: types 40 and 41) are a significant cause of gastroenteritis in infants and immunosuppressed populations. Our study found reduced IAP expression in infection with the *Adenovirus pan-genome*, consistent with prior evidence that adenoviruses use multiple immune evasion strategies ^41^. While a direct link between adenovirus and IAP suppression remains speculative, the presence of IAP inhibition in infected tissues indicates possible indirect modulation via inflammation.

Taken together, these findings support a hypothesis in which vitamin D status modulates IAP expression, which in turn influences vulnerability to enteric pathogens. IAP deficiency may serve as both a marker and a mediator of mucosal defence, and vitamin D deficiency further amplifies this vulnerability ^42^. The convergence of *E. bieneusi* and *Adenovirus* on this weakened defence network suggests a shared pathogenic strategy that exploits host nutrient and immune deficits. This hypothesis aligns with findings from studies linking low vitamin D to increased infection risk in hospitalized patients and supports the therapeutic potential of vitamin D and IAP-based interventions in reducing enteric disease burden ^43^. Vitamin D supplementation has been shown to enhance IAP activity, suggesting potential benefits in treating IAP-deficient states, including IBD and bacterial overgrowth.

This study contributes to the growing understanding of IAP and vitamin D as pivotal regulators of gastrointestinal health and systemic homeostasis. Their synergistic effects on gut microbiota, nutrient absorption, and immune modulation could offer a promising road for therapeutic intervention. While the associations presented are compelling, this study is observational and cannot establish causation. Moreover, most vitamin D and IAP studies have been performed in animal models, which lack the strong evidence in humans. Applied value may be achieved through further longitudinal trials that evaluate the effects of vitamin D and IAP supplementation on infection outcomes. By addressing the voids in our knowledge, future research can pave the way for innovative treatments targeting these crucial elements.

## Conclusion

IAP was found to be significantly and negatively associated with *Adenovirus-pan genome* and a less-explored enteric pathogen, *Enterocytozoon bieneusi*. IAP and vitamin D are integral to gastrointestinal health, with overlapping roles in nutrient absorption, microbiota regulation, and immune modulation. Further studies are required to determine and validate IAP’s therapeutic role against pathogenic infections in humans, exploring potential strategies for boosting IAP levels and elucidating the mechanisms underlying IAP and vitamin D interactions. Co-supplementation of vitamin D and IAP-boosting compounds, could be a novel approach for combating gastrointestinal and metabolic diseases.

## Contributions

JF conceptualized the study, analyzed the data, and drafted the initial manuscript. MAG contributed to data analysis, data interpretation and drafted the manuscript. MZI contributed to the analysis and interpretation of data, and drafted the manuscript. RAS was involved in data analysis, interpretation and drafting the manuscript. IM was involved in designing the study, methodology, reviewed and revised the manuscript, and supervised the work. MM conceptualized the study, and reviewed the manuscript. TA conceptualized the study, and reviewed the manuscript. JF is the guarantor of the study. All authors read and approved the final manuscript.

## Acknowledgement

The current manuscript did not receive any funding. However, we used data from the “MAL-ED (Etiology, Risk Factors, and Interactions of Enteric Infections and Malnutrition and the Consequences for Child Health and Development)” study, which was supported by the Bill & Melinda Gates Foundation. icddr,b is always grateful to it’s code donors, the Government of Bangladesh and the Government of Canada for their support. The funding sources had no influence on design of the study, data collection and analysis, or interpretation of the results.

## Ethical Standard Disclosure

All procedures were carried out in accordance with relevant regulations and the Institutional Review Board (IRB) of icddr,b, Mohakhali, Dhaka, Bangladesh had approved the research protocol (protocol number: 2008-020). Informed written consent were obtained from the caregivers of all the children at enrolment.

## Data Availability Statement

Data are available upon reasonable request. At icddr,b, we adhere to a strict policy to ensure that data containing identifying patient information is not made available. However, data related to this paper are available on request. Researchers who meet the criteria for accessing confidential data may request it by contacting Ms. Shiblee Sayeed from the Research Administration of icddr,b (http://www.icddrb.org/).

## Patient and Public Involvement

No patient or public were involved during any stage of the study.

